# Klotho plasma levels are an independent predictor of mortality in women with acute coronary syndrome

**DOI:** 10.1101/2024.11.01.24316615

**Authors:** Marcelino Cortés García, Andrea Kallmeyer Mayor, Nieves Tarín, Carmen Cristobal, Ana María Pello Lázaro, Alvaro Aceña, Carlos Gutierrez-Landaluce, Ana Huelmos, Joaquín Alonso Martín, Lorenzo López Bescós, Ignacio Mahillo-Fernandez, Oscar Lorenzo, Maria Gonzalez-Casaus, Jesus Egido, Jose Tuñon

## Abstract

**INTRODUCTION:** Alterations in plasma levels of the components of the mineral metabolism (MM) system (calcidiol, fibroblast growth factor-23 [FGF23], phosphate, parathormone [PTH] and klotho) are related to cardiovascular diseases. However, gender differences of the whole MM system in patients with acute coronary syndrome (ACS) have not been reported so far.

**METHODS:** We studied 1,230 patients with ACS. At baseline clinical data were collected and plasma levels of MM components were assessed. The primary outcome was a composite of acute ischaemic events, heart failure and all-cause mortality. Secondary outcomes included each component separately.

**RESULTS:** 282 patients (22.9%) were female. At baseline, FGF23, PTH, phosphate, and klotho plasma levels were higher in women than in men. After 5.44 (3.03-7.46) years of follow-up, the primary outcome occurred in 28.0% women and 23.5% men, and death in 10.6% and 9.4% respectively. At multivariate Cox regression analysis, FGF23 levels were associated with incidence of the primary outcome in both, women (Hazard ratio [HR] 1.02 [95%CI:1.01-1.04];p=0.001) and men (HR 1.04 [1.00-1.03];p=0.016), in whom calcidiol levels were also an independent predictor of this outcome. Klotho (HR 0.80 [95%CI:0.67-0.96];p=0.019) and phosphate (HR=2.24 [95%CI: 1.11-4.50];p=0.025) were independent predictors of death in women, while calcidiol (HR=0.84 [95%CI0.72-0.98];p=0.024) and FGF23 levels (HR=1.02 [1.00-1.03];p=0.048) were predictors in men.

**CONCLUSIONS:** Klotho levels are inversely and independently related to all-cause mortality after an ACS in women, but not in men. Furthermore, the MM profile in ACS patients differs in both genders. Future research should explore the underlying mechanisms of these associations.

## INTRODUCTION

The main role of the mineral metabolism (MM) system (calcidiol, fibroblast growth factor-23 [FGF23], phosphate, parathormone [PTH] and klotho) is to maintain mineral homeostasis. Specifically, FGF23 helps the failing kidneys to eliminate phosphorus and calcium (1). It also promotes a reduction of the concentration of 1-25-dihydroxyvitamin D, that leads to a decrease in intestinal calcium absorption, stimulating PTH secretion. However, these compensatory changes of the different components of MM may promote the development of cardiovascular disorders (CVD), and their plasma levels may also have a prognostic role in certain CVD (2–5). In the case of klotho, it works as a co-receptor of FGFR1 for the canonical actions of FGF23(6). The non-canonical actions of FGF23, such as cytokine production and the development of cardiac hypertrophy and fibrosis, are mediated mainly by FGFR2-4 receptors without the participation of klotho. Then, a decrease of klotho levels favours an increase of the deleterious, non-canonical effects of FGF23 (7), and it has been said that this molecule possesses protective effects (8–10).

In addition, these MM abnormalities are not limited to patients with chronic kidney disease (CKD), because they have been observed in patients with preserved renal function (11). Furthermore, a prognostic role of abnormal plasma levels of MM have been demonstrated in subjects with average renal function (4,12–14).

Moreover, CVD is the most frequent cause of mortality worldwide. In this setting, there are important gender differences in acute coronary syndromes (ACS): different comorbidities, cardiovascular risk factors, differences in clinical presentation and also in the quality of diagnostic and therapeutic medical management (15). Even more, the risk of death is known to be higher in women, especially in the context of younger populations (16–18).

To our knowledge, there are no previous publications in the literature assessing the prognostic role of the whole MM after an ACS focusing on gender differences. In this work we have analysed the potential differences on the prognostic role of MM in women suffering an ACS as compared to men.

## METHODS

### Patients and study design

We analysed the BACS & BAMI (Biomarkers in Acute Coronary Syndrome & Biomarkers in Acute Myocardial Infarction) study population, which included patients admitted to five hospitals in Madrid with ST-segment elevation myocardial infarction (STEMI) or non-ST-segment elevation acute coronary syndrome (NSTEACS). Inclusion and exclusion criteria have been detailed previously (19,20).Between July 2006 and June 2014, a total of 2,740 patients were discharged from the study hospitals with a diagnosis of NSTEACS or STEMI. Of these, 1,483 patients were excluded based on the following predefined criteria: presence of survival-limiting toxic conditions or habits (29.8%), age >85 years (16.4%),inability to complete follow-up (16.3%), clinical instability beyond day six after the index event (10.9%),inability of the investigators to include them (9.8%), inability to undergo cardiac revascularisation (9.6%), presence of other significant cardiac conditions (5.7%), and refusal to participate in the study (1.5%). Of the 1,257 patients included, 1,230 completed the follow-up.

On admission, baseline clinical variables were documented, and 12-hour fasting venous blood samples were collected in EDTA tubes. These samples were centrifuged at 2500 g for 10 minutes, and the plasma was stored at −80°C. After discharge from hospital, all patients underwent annual assessments at their respective medical centres. At the conclusion of the follow-up period, medical records were reviewed, and patient status was confirmed through telephone contact. The last follow-up visits were conducted in June 2016.

The research protocol suited the ethical guidelines of the1975 Declaration of Helsinki as reflected in a priori approval by the human research committees of the institutions participating in this study: Fundación Jiménez Díaz University Hospital, Fundación Alcorcón Hospital, Fuenlabrada Hospital, Puerta de Hierro Majadahonda University Hospital, and Móstoles University Hospital. All patients signed informed consent document. The date of approval by the Ethics Committee was 24 April 2007 (act number 05-07).

### Outcomes

The primary outcome was a composite of acute ischemic events (including non-STEMI, STEMI, unstable angina, stroke, and transient ischemic attack), heart failure, and all-cause mortality. Secondary outcomes included each component of the primary outcome: acute ischemic events, heart failure, and death. NSTEACS was defined as rest angina lasting more than 20 minutes within the preceding 24 hours, or new-onset class III-IV angina, accompanied by transient ST depression or T wave inversion on the electrocardiogram, as interpreted by the attending cardiologist, and/or elevated troponin levels. STEMI was defined by angina-like symptoms persisting for more than 20 minutes, ST elevation in at least two contiguous leads on the electrocardiogram, lack of response to nitroglycerin, and elevated troponin levels. A previous acute myocardial infarction was diagnosed in the presence of new pathological Q waves on the electrocardiogram, along with corresponding new myocardial scarring identified via echocardiography or nuclear magnetic resonance imaging. Stroke was defined as the rapid onset of a neurological deficit attributable to a specific vascular territory, lasting more than 24 hours, or confirmed by new ischemic lesions on imaging studies. A transient ischemic attack was characterized by transient neurological signs and symptoms of cerebral ischemia that resolved within 24 hours, without acute ischemic lesions on imaging studies.

Although all events were recorded for each patient, only the first event was included in the Cox regression analysis. Therefore, while the total number of events is reported, patients who experienced multiple events were counted only once in these analyses.

### Biochemical analysis

Plasma analyses were conducted at the Mineral Metabolism laboratory of La Paz Hospital and at the Vascular Pathology and Biochemistry laboratories of Fundación Jiménez Díaz University Hospital. The investigators responsible for these laboratory studies were blinded to the clinical data. Soluble-α-klotho levels (here in after referred to as “klotho”) were assessed by ELISA (Human Soluble Alpha Klotho Assay Kit, Immuno-Biological Laboratories Co., Hokkaido, Japan). FGF23 was measured through an enzyme-linked immunosorbent assay (ELISA) that targets epitopes within the caryboxyl-terminal region of FGF23 (Human FGF23, C-Term, Immutopics Inc, San Clemente, CA). Calcidiol plasma levels were quantified using a chemiluminescent immunoassay on the LIAISON XL analyzer (LIAISON 25OH-Vitamin D Total Assay, DiaSorin, Saluggia, Italy). Intact parathyroid hormone (PTH) was analyzed using a second-generation automated chemiluminescent method (Elecsys 2010 platform, Roche Diagnostics, Mannheim, Germany), and phosphate levels were determined by an enzymatic method (Integra 400 analyzer, Roche Diagnostics, Mannheim, Germany). N-terminal pro-brain natriuretic peptide (NT-proBNP) levels were measured via immunoassay (VITROS, Ortho Clinical Diagnostics, Raritan, NJ, USA), troponin through an immunometric immunoassay using a biotinylated monoclonal mouse antibody and a luminescent reaction (Ortho Clinical Diagnostics Vitros XT 7600, Raritan, NJ, USA), and high-sensitivity C-reactive protein (hs-CRP) by latex-enhanced immunoturbidimetry (ADVIA 2400 Chemistry System, Siemens, Munich, Germany). Lipid, glucose, and creatinine levels were determined using standard methods (ADVIA 2400 Chemistry System, Siemens, Munich, Germany). The estimated glomerular filtration rate (eGFR) was calculated using the Chronic Kidney Disease Epidemiology Collaboration (CKD-EPI) equation.

### Statistical analysis

Quantitative data following a normal distribution are presented as mean ± standard deviation, and those with a not normal distribution are displayed as median (interquartile range). Categorical variables are expressed using frequency measurements (absolute frequencies and percentages) A baseline comparative analysis of variables was conducted based on gender. Categorical data were evaluated using the χ² test or Fisher’s exact test. For continuous variables, a Student’s t-test was applied to those with a normal distribution, while the Mann–Whitney U test was employed for those not normally distributed. A p-value of less than 0.05 was considered indicative of statistical significance.

A univariate Cox regression analysis was conducted to determine which variables were associated with the development of various outcomes, separately for men and women. Subsequently, a multivariate regression analysis was performed to identify significant predictors of clinical outcomes in both genders. The selection criteria for variables included in the multivariate analysis were based on clinical and biological plausibility, as well as statistical significance observed in the univariate analyses. The magnitude of the effects of the variables was expressed in the form of hazard ratios (HRs) and 95% confidence intervals (CIs).

All analyses were conducted using the Statistical Package for the Social Sciences (SPSS v.26.0, IBM, Armonk, NY, USA), the R statistical language version 4.0.5 (R Foundation for Statistical Computing, Vienna, Austria) and the statistical package for the biomedical sciences (MedCalc v.23.0.2, Ostend, Belgium; https://www.medcalc.org).

## RESULTS

### Baseline characteristics

1,230 patients were included in our study. Of these, 282 (22.9%) were women. Women were older than men (65.7 vs 60.5 years, p<0.001), with a higher percentage of hypertension (70.6% vs 53.2%), and of CKD prior to admission (28.7% vs 15.9%) (Table 1). On the other hand, men had a significantly higher rate of smokers (46.6% vs 28.0%), with more coronary heart disease and peripheral arterial disease prior to inclusion in the study. In both genders, the percentage of STEMI included in our population was around 50%. On average, men had a greater number of affected vessels (1.51 vs. 1.22, p<0.001), and a higher percentage of them received revascularization treatment. This translated into a lower proportion of women with P2Y12 inhibitors at discharge (85.5% vs 91.6%, p 0.004), with no other difference in treatment at discharge between men and women. Regarding the biomarkers analyzed, women generally presented significantly higher levels of PTH, FGF23, klotho, phosphorus, and NT-proBNP. Median time for blood extraction from admission was 4 (2–5) days.

**Table 1:** Baseline characteristis of study population.

| Variable | Female(n=282) | Male (n=948) | p |
| --- | --- | --- | --- |
| Age (years) | 65.7 ± 13.0 | 60.5 ± 11.6 | <0.001 |
| Age > 70 years | 127 (45.0%) | 241 (25.4%) | <0.001 |
| Caucasian race | 275 (97.5%) | 917 (96.7%) | NS |
| Diabetes | 70 (24.8%) | 210 (22.2%) | NS |
| Hypertension | 199 (70.6%) | 504 (53.2%) | <0.001 |
| Body mass index (kg/m<sup>2</sup>) | 28.5 ± 5.16 | 28.3 ± 4.02 | NS |
| Dyslipidaemia | 165 (58.5%) | 575 (60.7%) | NS |
| Smoking | 79 (28.0%) | 442 (46.6%) | <0.001 |
| eGFR < 60 mL/min/1.73 m<sup>2</sup> | 81 (28.7) | 151 (15.9) | < 0.001 |
| Heart failure (prior to inclusion) | 4 (1.4%) | 10 (1.1%) | NS |
| Coronary artery disease (prior) | 42 (14.9%) | 202 (21.3%) | 0.022 |
| Peripheral artery disease (prior) | 7 (2.5%) | 64 (6.8%) | 0.011 |
| Cerebrovascular accident (prior) | 13 (4.6%) | 27 (2.8%) | NS |
| Atrial fibrillation (prior) | 8 (2.8%) | 20 (2.1%) | NS |
| **Laboratory results** |  |  |  |
| Hemoglobin (g/dl) | 13.5 ± 1.50 | 15.1 ± 1.88 | <0.001 |
| Glycaemia (mg/dL) | 116 ± 38.4 | 117 ± 39.1 | NS |
| eGFR (mL/min/1.73 m<sup>2</sup>) | 74.9 ± 22.3 | 79.8 ± 19.2 | 0.001 |
| Troponin (ng/mL) | 8.13 (0.80, 30.1) | 11.4 (0.82, 59.4) | 0.022 |
| LDL (mg/dL) | 119 ± 39.4 | 116 ± 36.1 | NS |
| Triglycerides (mg/dL) | 122 (82.5, 177) | 134 (98.0, 185) | 0.006 |
| HDL (mg/dL) | 47.2 ± 14.2 | 38.1 ± 10.2 | <0.001 |
| Calcidiol (ng/mL) | 16.4 (12.4, 24.6) | 18.6 (13.5, 24.7) | 0.095 |
| PTH (pg/mL) | 54.0 (40.3, 70.0) | 45.0 (36.0, 60.0) | <0.001 |
| FGF23 (RU/mL) | 126 (95.0, 173) | 105 (82.0, 138) | <0.001 |
| Klotho (pg/mL) | 657 ± 219 | 621 ± 205 | 0.013 |
| Phosphorus (mg/dL) | 3.44 (3.12, 3.82) | 3.21 (2.85, 3.58) | <0.001 |
| NT-proBNP (pg/mL) | 551 (226, 1500) | 348 (121, 912) | <0.001 |
| hs-CRP (ng/dL) | 1.36 (0.59, 2.85) | 1.74 (0.78, 3.62) | 0.012 |
| **Type of ACS and coronary angiographical findings** |  |  |  |
| NSTEMI | 114 (40.4%) | 326 (34.4%) | NS |
| STEMI | 129 (45.7%) | 482 (50.8%) | NS |
| Unstable Angina | 39 (13.8%) | 140 (14.8%) | NS |
| LVEF < 40% | 41 (14.6%) | 131 (13.8%) | NS |
| Number of affected vessels | 1.22 ± 0.80 | 1.51 ± 0.79 | <0.001 |
| Left main disease | 9 (3.3%) | 33 (3.5%) | NS |
| Complete revascularization | 211 (74.8%) | 660 (69.6%) | NS |
| **Type of revascularization** |  |  | <0.001 |
| No revascularization | 70 (24.8%) | 115 (12.1%) |  |
| Drug eluting stent | 142 (50.4%) | 492 (51.9%) |  |
| Bare metal stent | 52 (18.4%) | 261 (27.5%) |  |
| Balloon angioplasty | 11 (3.9%) | 27 (2.8%) |  |
| Coronary artery bypass grafting | 7 (2.5%) | 53 (5.6%) |  |
| **Treatments at discharge** |  |  |  |
| ASA | 264 (93.6%) | 911 (96.2%) | NS |
| P2Y12 inhibitors | 241 (85.5%) | 867 (91.6%) | 0.004 |
| Anticoagulant | 22 (7.8%) | 60 (6.3%) | NS |
| Statins | 264 (93.6%) | 916 (96.7%) | NS |
| Ezetimibe | 4 (1.4%) | 18 (1.9%) | NS |
| Insulin | 25 (8.9%) | 57 (6.0%) | NS |
| Oral antidiabetic drugs | 39 (13.8%) | 149 (15.7%) | NS |
| ACEI/ARB | 227 (80.5%) | 770 (81.3%) | NS |
| MRAs | 26 (9.2%) | 71 (7.5%) | NS |
| Beta-blockers | 227 (80.5%) | 789 (83.4%) | NS |
| Nitrates | 53 (18.8%) | 133 (14.0%) | NS |
| Diuretics | 48 (17.0%) | 139 (14.7%) | NS |

ACEI: angiotensin converting enzyme inhibitor; ARB: angiotensin receptor blocker; ASA: acetylsalicylic acid; eGFR: estimated glomerular filtration rate; FGF23: fibroblast growth factor 23; HDL: high density lipoprotein; hs-CRP: high sensitivity C-reactive protein; LDL: low density lipoprotein; LVEF: left ventricular ejection fraction; NS: statistically not significant; NSTEMI: non-ST elevation myocardial infarction; MRAs: mineralocorticoid receptor antagonists; NT-ProBNP: N-terminal-probrainnatriuretic peptide; PTH: parathormone; STEMI: ST-elevation myocardial infarction.

### Primary outcome

Median follow-up was 5.44 (3.03-7.46) years. During follow-up, 79 women (28.0%) and 223 men (23.5%) developed a primary event (a composite of acute ischemic events, heart failure, and all-cause mortality).

We performed a multivariate Cox regression analysis of our study population to identify independent predictors of primary outcomes separately for men and women, as described previously. This analysis revealed that FGF23 levels were directly and significantly related to the occurrence of a primary outcome in both sexes (Table 2). PTH also showed a relationship with primary outcome in the male group, but not in the female group. Other variables (clinical or treatment) also showed a significant relationship with outcomes in both groups, although no other biomarker showed a significant relationship.

**Table 2:** Multivariable Cox regression analysis for the primary outcome of acute ischemic event, heart failure or death.

| **Gender** | **Variable** | **HR (CI 95%)** | **p** |
| --- | --- | --- | --- |
| **Female** | eGFR | 0.85 (0.77-0.95) | 0.006 |
|  | Nitrates | 2.34 (1.40-3.90) | 0.001 |
|  | CVA | 4.17 (1.98-8.78) | <0.001 |
|  | FGF23 | 1.02 (1.01-1.04) | 0.001 |
|  | Diltiazem | 2.41 (1.07-5.40) | 0.034 |
|  | Diabetes | 1.65 (1.02-2.66) | 0.040 |
|  | Statins | 0.497 (0.25-0.99) | 0.049 |
| **Male** | Age | 1.0 (1.02-1.04) | <0.001 |
|  | Diuretics | 1.77 (1.29-2.43) | <0.001 |
|  | CAD | 1.40 (1.05-1.88) | 0.023 |
|  | Diabetes | 1.50 (1.13-2.01) | 0.006 |
|  | ASA | 0.52 (0.31-0.86) | 0.012 |
|  | PTH | 1.06 (1.01-1.11) | 0.016 |
|  | FGF23 | 1.04 (1.00-1.03) | 0.027 |

ASA: acetylsalicylic acid; CVA: cerebrovascular accident (prior to inclusion); CAD: coronary artery disease (prior to inclusion); eGFR: estimated glomerular filtration rate; FGF23: fibroblast growth factor 23; HR: Hazard Ratio; PTH: parathormone.

### Secondary outcomes

At the end of follow-up, 55 women (19.5%) and 133 men (14%) presented an acute ischemic event. For each of these groups, we performed a multivariate Cox regression analysis to identify significant predictors of this outcome (Table 3). Variables such as eGFR, heart failure (prior to inclusion), hypertension, etc. were shown to be a predictor of acute ischemic events. However, none of the analyzed biomarkers showed a statistically significant relationship in either group.

**Table 3:** Multivariable Cox regression analysis for the secondary outcome of acute ischemic events.

| **Gender** | **Variable** | **HR (CI 95%)** | **p** |
| --- | --- | --- | --- |
| **Female** | Heart failure | 6.43 (2.09-19.79) | 0.002 |
|  | Nitrates | 3.19 (1.79-5.66) | <0.001 |
|  | Glycaemia | 1.09 (1.03-1.15) | 0.004 |
|  | eGFR | 0.87 (0.76-0.98) | 0.028 |
| **Male** | CCB | 2.43 (1.59-3.40) | <0.001 |
|  | Heart failure | 3.75 (1.37-10.29) | 0.011 |
|  | Hypertension | 1.75 (1.20-2.57) | 0.004 |
|  | Triglycerides | 1.01 (1.00-1.02) | 0.018 |
|  | ACEI/ARB | 0.65 (0.44-0.97) | 0.033 |

ACEI: angiotensin converting enzyme inhibitor; ARB: angiotensin receptor blocker; CCB: calcium channel blocker; eGFR: estimated glomerular filtration rate; HR: Hazard Ratio.

Twenty women (7.1%) and 48 men (5.1%) developed heart failure during follow-up. After multivariate Cox regression analysis, again FGF23 was an independent risk factor for the development of heart failure in both men and women (Table 4). PTH also showed a statistically significant relationship with this event but, as observed for the primary outcome, only in the male population.

**Table 4:** Multivariable Cox regression analysis for the secondary outcome of heart failure.

| **Gender** | **Variable** | **HR (CI 95%)** | **p** |
| --- | --- | --- | --- |
| **Female** | MRAs | 9.22 (3.21-26.55) | <0.001 |
|  | FGF23 | 1.03 (1.01-1.06) | 0.009 |
|  | Insulin | 5.56 (1.71-18.31) | 0.008 |
|  | ASA | 0.23 (0.06-0.94) | 0.042 |
| **Male** | FGF 23 | 1.04 (1.03-1.06) | <0.001 |
|  | PTH | 1.14 (1.06-1.23) | 0.002 |
|  | Age | 1.06 (1.02-1.09) | 0.001 |
|  | Oral antidiabetic drugs | 3.02 (1.60-5.71) | 0.001 |
|  | LVEF < 40% | 3.73 (1.99-6.95) | <0.001 |
|  | Diltiazem | 4.97 (1.68-14.69) | 0.005 |
|  | Hypertension | 2.23 (1.03-4.84) | 0.043 |

ASA: acetylsalicylic acid; eGFR: estimated glomerular filtration rate; FGF23: fibroblast growth factor 23; LVEF: left ventricular ejection fraction; MRAs: mineralocorticoid receptor antagonists; HR: Hazard Ratio; PTH: parathormone

Finally, we analyzed the variables related to all-cause mortality in both genders. By the end of follow-up, 30 women (10.6%) and 89 men (9.4%) had died. We observed that klotho was an independent protective factor in the female population for all-cause mortality (HR 0.80, CI95% 0.67-0.96; p=0.019) (Table 5). Phosphate levels were also independently but positively associated with this outcome in this population (HR 2.24 (1.11-4.50; p=0.025). In contrast, in men the MM components associated with this outcome included only FGF23 (HR 1.02 (1.00-1.03); p=0.048) and calcidiol (HR 0.84 (0.72-0.98); p=0.024) but not klotho and phosphate plasma levels.

**Table 5:** Multivariable Cox regression analysis for the secondary outcome of all-cause mortality.

| **Gender** | **Variable** | **HR (CI 95%)** | **p** |
| --- | --- | --- | --- |
| **Female** | eGFR | 0.97 (0.95-0.98) | 0.001 |
|  | Anticoagulant | 6.77 (2.69-17.05) | <0.001 |
|  | Diuretics | 3.67 (1.58-8.51) | 0.004 |
|  | P2Y12 inhibitors | 0.33 (0.14-0.77) | 0.013 |
|  | Phosphorus | 2.24 (1.11-4.50) | 0.025 |
|  | Klotho | 0.80 (0.67-0.96) | 0.019 |
|  | LVEF < 40% | 2.45 (1.05-5.76) | 0.040 |
| **Male** | Age | 1.10 (1.07-1.13) | <0.001 |
|  | Insulin | 2.59 (1.44-4.66) | 0.002 |
|  | NT-proBNP | 1.02 (1.01-1.03) | 0.002 |
|  | Heart failure (prior) | 4.28 (1.78-10.27) | 0.001 |
|  | ASA | 0.41 (0.20-0.84) | 0.015 |
|  | Smoking | 1.71 (1.00-2.92) | 0.049 |
|  | Calcidiol | 0.84 (0.72-0.98) | 0.024 |
|  | FGF23 | 1.02 (1.00-1.03) | 0.048 |

ASA: acetylsalicylic acid; eGFR: estimated glomerular filtration rate; FGF23: fibroblast growth factor 23; LVEF: left ventricular ejection fraction; HR: Hazard Ratio; NT-ProBNP: N-terminal-probrain natriuretic peptide.

## DISCUSSION

Many differences have been described between men and women regarding coronary artery disease. Women have less coronary atherosclerosis, (21) and a lower risk of suffering ACS than men (22). In addition, the age of onset is higher in women than in men, with a higher prevalence of comorbidities (23). The impact of the different risk factors according to gender also presents differences; with the greater importance in women of diabetes, hypertension and smoking (24–26). Thus, the risk of death is known to be higher in women. This higher mortality in women is described mainly in the earlier phases after ACS, with a worse prognosis and higher mortality having been observed immediately after percutaneous intervention(23,27). The possible causes of this worse prognosis are diverse, and they may include a delay in the diagnosis of ACS favored by the higher likelihood of atypical symptoms(28)and differences in high-sensitivity troponin levels as compared to men (29). Furthermore, women with ACS are less likely to receive optimal therapy than men (30–34).

Changes in plasma levels of the different components of MM have been related to different cardiovascular abnormalities. Low calcidiol and high PTH levels have been associated with left ventricular hypertrophy, hypertension, coronary heart disease and increased cardiovascular risk (2,20,35). Increased FGF23 has been related with left ventricular hypertrophy (3,36,37), heart failure (38,39), atrial fibrillation (40) and coronary heart disease (4)and is associated with worse prognoses both in the general population and in patients with CVD (5,41–44). Furthermore, MM abnormalities are not restricted to patients with CKD, but they are also present in patients with normal renal function(11), where they may also have prognostic value(4,12–14).In spite of this, there is no information regarding the behaviour of MM components in women with ACS.

In this paper we demonstrate that women with ACS present a worse MM profile than men, with higher FGF23, PTH, and phosphate levels. Regarding prognosis, FGF23 levels were independent predictors of the primary outcome in both genders, while PTH added independent predictive value only in men. Similar results were obtained in the prediction of heart failure. However, for the prediction of all-cause mortality there were marked differences between both groups, with low klotho levels being predictive in women, along with phosphate levels, while in men high FGF23 and low calcidiol levels were the independent predictors for this outcome.

Klotho is the co-receptor of FGFR1 for FGF23, which exerts its beneficial actions through this canonical pathway, such as helping the failing kidneys to eliminate phosphate. However, in the absence of klotho, FGF23 binds to other receptors promoting cardiac hypertrophy and fibrosis and stimulating the production of pro-inflammatory cytokines(7). Then, it has been said to have protective effects. Klotho suppression in animal models is associated with early ageing, shorter life expectancy, multi-organ dysfunction and marked alterations in mineral homeostasis (hyperphosphataemia, hypercalcaemia, among others.)(45,46). More recently, an independent association between low klotho levels with left ventricular hypertrophy (47), heart failure(48–51), atrial fibrillation (52) and myocardial infarction (49)has been described in humans. Klotho has also demonstrated a prognostic role in heart failure(48,53), with low levels also being a marker of total and cardiovascular mortality in CKD patients(54) and in the general population (55,56).

Basic research studies report results that may explain in part a potential benefit of klotho in ACS. The administration of klotho in animal models protects the myocardium from ischaemic and reperfusion damage through the activation of different pathways that reduce oxidative stress and restore autophagy levels (57,58). Klotho has been described to modulate platelet activity (59). Klotho therapy improves cardiac remodelling in a murine model of myocardial infarction (60) and it also ameliorates diastolic function (61), although there are not clinical studies confirming these findings. In several other studies, authors have also observed a beneficial effect of klotho on the vascular endothelium, reducing oxidative stress at this level or attenuating cell apoptosis, among other mechanisms (62–65).

However, there are no published data on the prognostic role of klotho after an ACS. To our knowledge, our work is the first to describe an independent protective role of klotho after ACS. According to the present findings, we have recently shown that cardiac rehabilitation after ACS is associated with increases of klotho levels (66), suggesting an increase in klotho could explain, at least in part, the benefits of cardiac rehabilitation. Of interest, the protective role of klotho is limited to women. It is possible that the higher cardiovascular risk profile of men, with more extensive coronary disease and a greater number of affected vessels, may interfere with the prognostic role of klotho, attenuating its protective effect after ACS. It is also striking that our results show that other MM biomarkers have a prognostic value in men but not in women. Nevertheless, we have demonstrated previously that it is common that several MM biomarkers have independent prognostic value (14,41), even after adjusting for established biomarkers such as NT-proBNP. This underlines the need to make a complete assessment of MM to investigate the prognostic value of its components.

### Limitations

First, the percentage of women in our study population is relatively low. This is in line with many other published studies. There are several factors that may influence the lower recruitment of women, among them, a higher prevalence of ACS in men could partly account for this difference. Second, the design of the study required the collection of plasma for analysis at discharge no later than 6 days after admission, to achieve homogeneous results. This led to the exclusion of ACS patients who did not meet this condition and that fact may explain the low number of cases with LVEF *<*40% that were included. Therefore, these results should not be extrapolated to populations with a high percentage of patients with moderate or severe LV systolic dysfunction.

## Conclusions

In conclusion, our results show that klotho levels are inversely related to all-cause mortality in women after an ACS, highlighting a possible gender-specific prognostic biomarker. These results underline the importance of considering MM biomarkers in the risk stratification and management of ACS patients, with attention to gender differences. Future research should explore the underlying mechanisms of these associations.

## Data Availability

All data referred to in the manuscript are available upon reasonable request to the corresponding author (Dr. Marcelino Cortés).

## Funding support/Acknowledgements

This work was supported by grants from Carlos III Health Institute (ISCIII) (PI17/01495; PI20/00923; PI23/00119; PI24/00978), Spain’s Ministry of Science and Innovation (RTC2019-006826-1), Spanish Society of Cardiology and Carlos III Health Institute FEDER (FJD biobank: RD09/0076/00101).

## Notes

### Competing Interest Statement

The authors have declared no competing interest.

### Clinical Trial

Our prospective study did not involve any specific intervention or treatment. However, the research protocol suited the ethical guidelines of the1975 Declaration of Helsinki as reflected in a priori approval by the human research committees of the institutions participating in this study: Fundación Jiménez Díaz University Hospital, Fundación Alcorcón Hospital, Fuenlabrada Hospital, Puerta de Hierro Majadahonda University Hospital, and Móstoles University Hospital. The date of approval by the Ethics Committee was 24 April 2007 (act number 05-07).

### Author Declarations

The protocol was approval by the human research committees of the institutions participating in this study: Fundación Jiménez Díaz University Hospital, Fundación Alcorcón Hospital, Fuenlabrada Hospital, Puerta de Hierro Majadahonda University Hospital, and Móstoles University Hospital. The date of approval by the Ethics Committee was 24 April 2007 (act number 05-07).

